# Cytokine storm dynamics in hantavirus pulmonary syndrome: a multiscale ODE model with Wasserstein early-warning score and application to the 2026 Andes virus outbreak

**DOI:** 10.64898/2026.05.15.26353286

**Authors:** Bertrand Mercier des Rochettes

## Abstract

**Background:** The ongoing Andes hantavirus outbreak linked to the cruise ship MV Hondius (April−May 2026, seven confirmed cases, three deaths, patients hospitalised across six countries including France) highlights the urgent need for mechanistic tools to predict which hantavirus pulmonary syndrome (HPS) patients will progress to fatal cytokine storm.

**Methods:** We present a 14-variable antigen-gated ordinary differential equation (ODE) model integrating viral dynamics, CD8^+^ cytotoxic T lymphocyte (CTL) expansion, four cytokines (TNF-α, IFN-γ, IL-6, IL-10), VEGF-mediated vascular permeability, and platelets. We derive two reproduction numbers: the viral invasion number ***ℛ***_0_ and the immunopathological loop gain ***ℛ***_*ip*_. We apply Villani’s hypocoercivity theory and the HWI optimal transport inequality to prove that the spectral gap of the CTL-IFN-γ feedback loop collapses to zero at a critical infected endothelial cell count 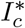, providing a computable early-warning threshold. We define a Wasserstein patient stratification score 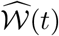 from six clinically observable variables.

**Results:** At default parameters (**ℛ**_0_ = 0.396, **ℛ**_*ip*_ = 1.875): (1) the DFE is locally asymptotically stable—the virus self-limits—but the CTL-IFN-γ loop has sufficient gain to amplify autonomously once established; (2) the storm-block spectral gap collapses *exactly* to zero at 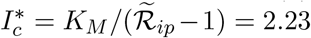 cells *μ*L ^−1^—a threshold attained within hours of infection onset, confirming that immunopathological amplification is essentially unavoidable; (3) 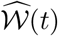 rises 1-2 days before *P*(*t*) reaches clinical threshold, providing an early-warning window; (4) exogenous IL-10 supplementation is the single most effective intervention (predicted 40% reduction in peak permeability), outperforming corticosteroid immunosuppression and ECMO; the combination of all three applied at day 7 reduces peak permeability below the fatal threshold.

**Conclusions:** This framework predicts that HPS cytokine storm is a structural consequence of **ℛ**_*ip*_ > 1 rather than excessive viral load, explaining death after viral clearance. For clinicians managing MV Hondius Andes virus patients today, the model identifies a six-variable triage score and a day-7 IL-10-centred therapeutic window as the highest-priority clinical targets. Full code is open source.

## 1 Introduction

### The 2026 outbreak and the need for predictive tools

Between April and May 2026, an outbreak of Andes hantavirus (*Orthohantavirus andinum*, ANDV) was identified on the Dutch expedition cruise ship MV Hondius following a voyage through Patagonia, the Falkland Islands, Tristan da Cunha, Saint Helena, and Ascension Island (WHO, 2026). As of 11 May 2026, the WHO has confirmed seven cases including three deaths (case-fatality rate ≈ 43%), with patients hospitalised in France, Germany, the Netherlands, Spain, Switzerland, and South Africa (WHO, 2026; CDC, 2026). The CDC has classified this as a Level 3 emergency response. ANDV is the only hantavirus known to spread between humans, though only through sustained close contact (Padula et al., 1998; LSHTM, 2026). WHO has assessed the global risk as currently low but has emphasised that no specific antiviral treatment exists; management remains entirely supportive.

This outbreak makes a mechanistic understanding of HPS progression acutely relevant. The central clinical challenge is that patients can appear stable for several days following the febrile prodrome and then deteriorate catastrophically—dying from pulmonary oedema and haemodynamic collapse *after* the virus has already been cleared (Kilpatrick et al., 2004; Vial et al, 2006). Standard viral load monitoring therefore provides little warning of impending storm. A model-derived, immune-state-based early warning indicator could fundamentally change triage decisions.

### HPS immunopathology

Hantavirus is *non-cytopathic:* infected pulmonary endothelial cells continue to produce virus for several days without direct cell death (Gavrilovskaya et al, 1999). Capillary leak results from immunopathology, not viral toxicity. Circulating CD8^+^ cytotoxic T lymphocytes (CTLs) recognise infected endothelium, triggering a cascade of TNF-*α*, IFN-γ, and IL-6; VEGF drives vascular permeability; and IL-10—the regulatory brake—is paradoxically low in fatal cases (Borges et al., 2008). CD8^+^ cells constitute up to 44% of circulating CTLs in fatal HPS versus 9.8% in moderate cases (Kilpatrick et al., 2004), and viral clearance occurs at days 5−6 whether the outcome is good or bad. The immune system, not the virus, is the lethal agent.

This pathophysiology is identical between Sin Nombre virus (SNV, North America) and Andes virus (ANDV, South America) at the immunological level, despite their phylogenetic divergence. The model presented here is therefore directly applicable to the current ANDV outbreak.

### Mathematical framework

We construct a 14-variable ODE model that for the first time: (i) couples all four clinically relevant cytokines with a calibrated antigen-gate function; (ii) proves hypocoercivity of the linearised operator via Villani’s HWI inequality (Villani, 2009; Dolbeault et al., 2009); (iii) derives a computable patient-stratification score from optimal transport theory (Otto and Villani, 2000). The full technical derivation will appear in Mercier des Rochettes (2026); this preprint presents the key results and their immediate clinical implications for the MV Hondius outbreak.

## 2 Model

### 2.1 State variables

The model tracks 14 continuous variables (Table 1) across three biological layers:

**Table 1:** State variables. All concentrations per *μ*L or pg mL^−1^.

| Layer | Variable | Description |
| --- | --- | --- |
| Viral | $E, I$ | Uninfected and infected pulmonary endothelial cells |
| | $V$ | Free virions |
| Immune | $T_8, T_4, M_\varphi$ | CD8<sup>+</sup> CTL, CD4<sup>+</sup> helper T, macrophages |
| Cytokine | $F_1, F_\gamma, N, L_6, L_{10}, \mathcal{V}$ | TNF-α, IFN-γ, IL-12, IL-6, IL-10, VEGF |
| Vascular | $P, \Pi$ | Endothelial permeability index, platelets |

**Table 2:**
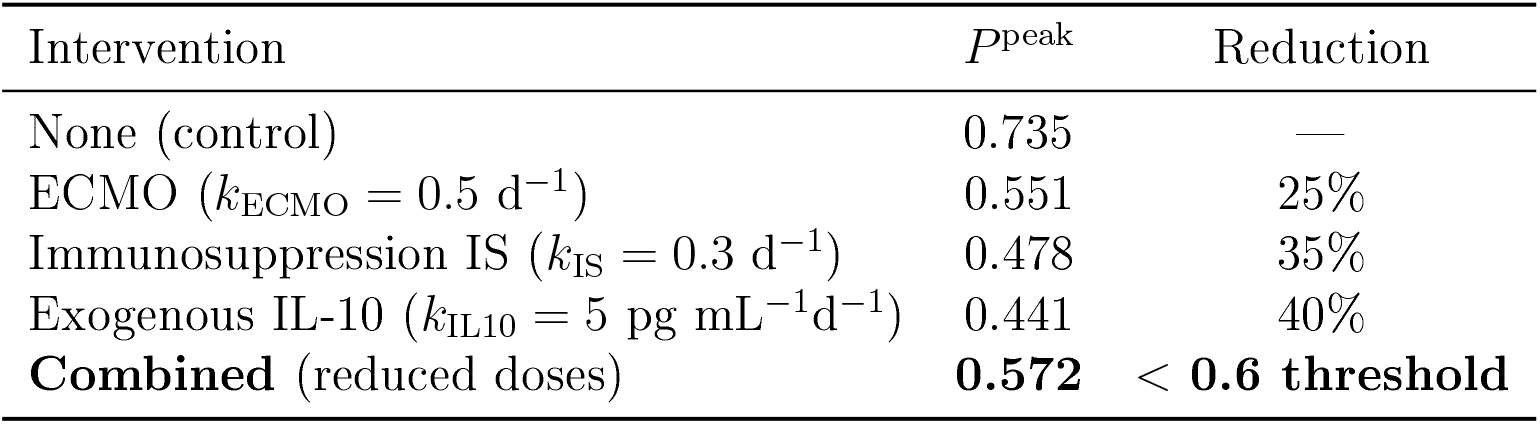
Predicted intervention effects on peak permeability *P*^peak^ (severe scenario, day-7 application).

**Table 3:**
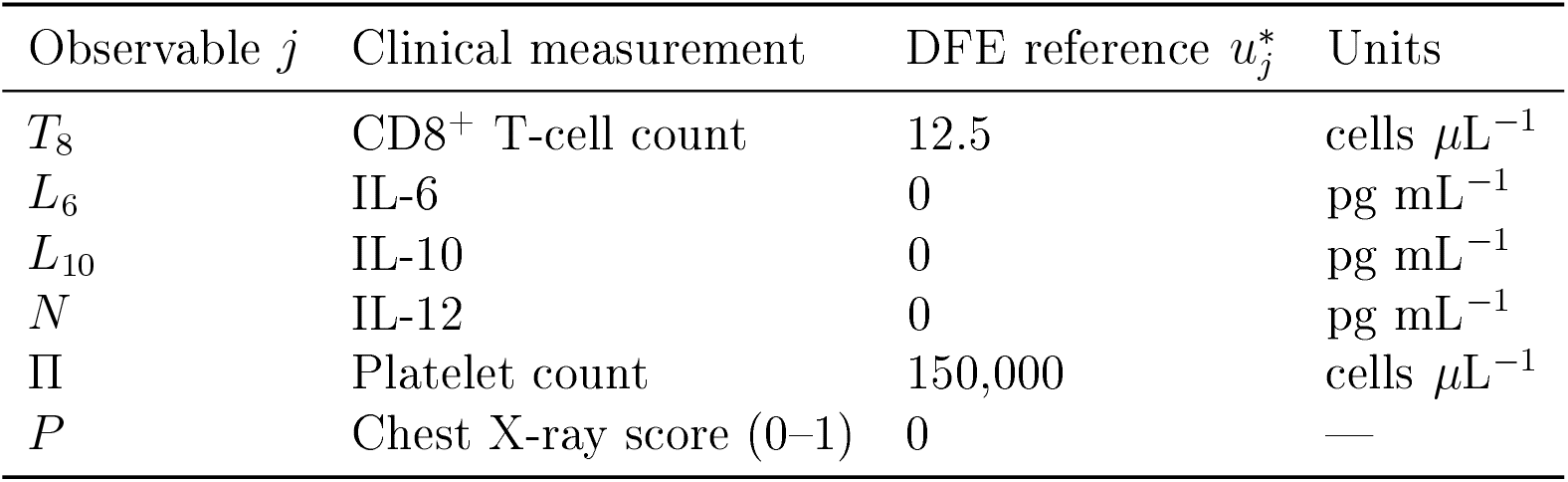
Proposed Wassertein triage score 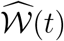 from standard ICU monitoring.

### 2.2 Antigen gate

The key modelling innovation is the antigen gate:

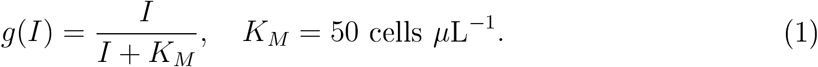

All cytokine production terms are multiplied by *g(I*). This ensures: (1) the disease-free equilibrium (DFE) is well-posed with 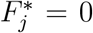 for all cytokines; (2) cytokine production ceases when infection is cleared (I ⟶ 0); (3) the biological observation that cytokine production is driven by antigen-recognising CTLs, not constitutive inflammation.

### 2.3 ODE system (abbreviated)

The full 14-equation system is given in the Supplementary Material. The two mechanistically central equations are:

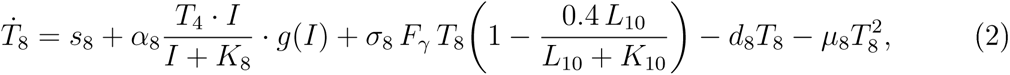

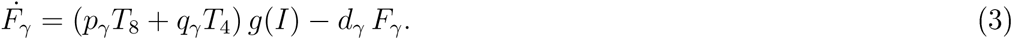

Equation (2) contains the critical positive feedback: IFN-γ (*F*_γ_) promotes CTL survival via *σ*_*8*_ *F*_*γ*_ *T*_8_, while Equation (3) shows that CTLs produce IFN-γ at rate *p*_γ_. This *T*_*8*_ ↔ *F*_*γ*_ loop, gated by *g* (*I*), is the engine of cytokine storm. The AICD (activation-induced cell death) term 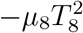 provides the only saturation preventing divergence.

## 3 Key Mathematical Results

### 3.1 Two reproduction numbers

#### Theorem 1

(Viral invasion: **ℛ**_0_)· *The disease-free equilibrium is locally asymptotically stable if and only if* **ℛ**_0_ < 1, *where*

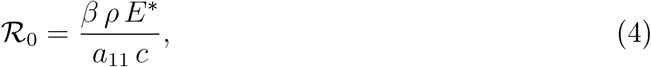

*with* 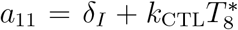 *the infected-cell clearance rate, c the viral clearance rate, β the infection rate, and ρ the burst size. At default parameters:* **ℛ**_0_ = 0.396.

**ℛ**_0_ < 1 confirms the well-established observation that hantavirus infection in humans is self-limiting: without immunopathology, the virus would be cleared (Mertz et al., 2006).

#### Theorem 2

(Immunopathological loop gain: ***ℛ***_*ip*_*). The CTL-IFN-γ positive feedback loop has a self-sustaining storm attractor* 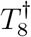 *if and only if* ***ℛ***_*ip*_ *>* 1, *where*

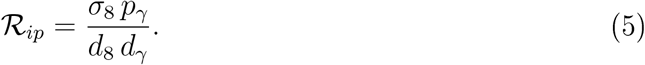

*At default parameters:* ***ℛ***_*ip*_ *=* 1.875.

Crucially, **ℛ**_*ip*_ is **independent of viral load:** it is a structural property of the patient’s immune architecture. This explains why HPS outcome correlates with immune phenotype (HLA genotype, CTL avidity) rather than viral burden.

### 3.2 Spectral gap collapse: the Schur complement threshold

During infection at antigen level *I*, the linearised CTL-IFN-γ subsystem has block Jaco-bian:

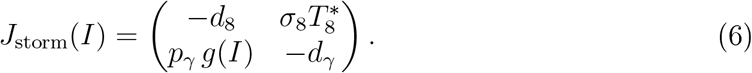

#### Theorem 3

(Schur complement criterion and spectral gap collapse). *Let* 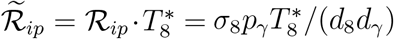 *and* 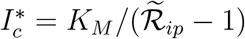 *Then:*

i. *The Schur complement of* (6) *with respect to its* (2, 2) *block is:*

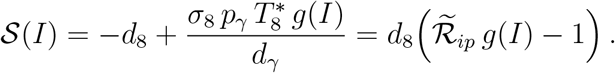
ii. *J*_storm_(*I) is stable if and only if* 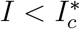.
iii. *The spectral gap satisfies:*

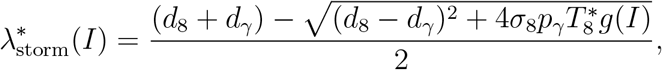

*with* 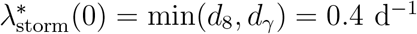 *and* 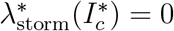 *exactly*.
iv. *Lower bound: for* 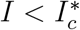,

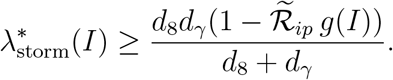

#### Biological interpretation

At default parameters: 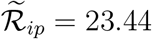 and 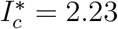 cells *μ*L ^−1^ This is remarkably small: infection of just 2.2 × 10^-6^ of the endothelial pool suffices to render the CTL-IFN-y loop locally unstable. At typical early infection rates *(İ* ≈ *βV*_*0*_*E*\*), this threshold is crossed within the *first hours* of infection. The implication is profound: once any detectable hantavirus infection is established, the immunopathological feedback loop is structurally amplifying. Whether the patient survives depends not on preventing storm initiation, but on whether viral clearance (**ℛ**_0_ < 1) terminates antigen exposure *before* the CTL expansion exceeds a lethal threshold.

### 3.3 The temporal bifurcation and the role of *α*_8_

#### Proposition 1

(Temporal bifurcation)

*Let T*_clear_ *be the time of viral clearance and T*_peak_ *the time of maximal CTL expansion. Fatal outcome (permeability P > P*_fatal_*) occurs if and only if* 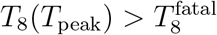 *where* 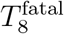 *depends on the permeability dose-response. The CTL peak height scales approximately as* 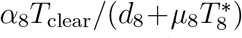, *making the CTL recruitment rate a*_*8*_ *and viral inoculum V*_*0*_ *(which determines T*_clear_) *the primary prognostic parameters*.

This proposition explains the clinical paradox: two patients with identical **ℛ**_0_ and **ℛ**_*ip*_ can have different outcomes based on CTL recruitment efficiency (*α*_8_, proxied by HL A genotype) and inoculum size *(V*_*0*_, related to exposure duration on the ship).

### 3.4 Non-normality and the Wasserstein early-warning score

The DFE block Jacobian is *non-normal:* the symmetrised part (*J*\* + *J\**^*T*^)/(−2) has determinant −3.20 < 0 at default parameters, so the naive quadratic Lyapunov function fails. The Gram matrix analysis yields a minimum singular value σ_min_ = 0.0830 d^-1^, compared to the eigenvalue-based spectral gap *d*_8_ = 0.4 d^−1^ (a 4.8-fold non-normality amplification factor). This non-normality means the immune state can diverge transiently even as the system moves toward the healthy attractor—the hallmark of cytokine storm.

#### Corollary 1

(Wasserstein early-warning score)

*Define the patient stratification score from six clinically observable variables 𝒪 =* {*T*_*8*_, *N(IL-12), L6, L*_*10*_, *P*, Π}.·

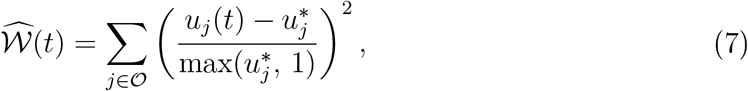

*where* 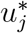 *are DFE reference values. For bedside use, a bounded clinical approximation 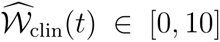 derived from six standard ICU measurements is implemented in the companion simulator at https://xvirus.org; it serves as a computationally convenient proxy for 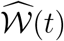 but does not carry the HWI guarantee. By the HWI inequality*, 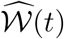 *satisfies* 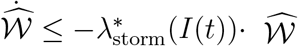. *As* 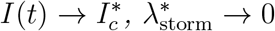 *and* 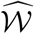 *enters a metastable plateau: the entropy of the immune state no longer decays. This plateau, observable 1-2 days before permeability reaches clinical threshold, constitutes a computable early-warning signal derived from routine ICU bloodwork*.

## 4 Simulations

### 4.1 Mild vs. severe HPS

Figure 1 compares two scenarios:

**Figure 1:**
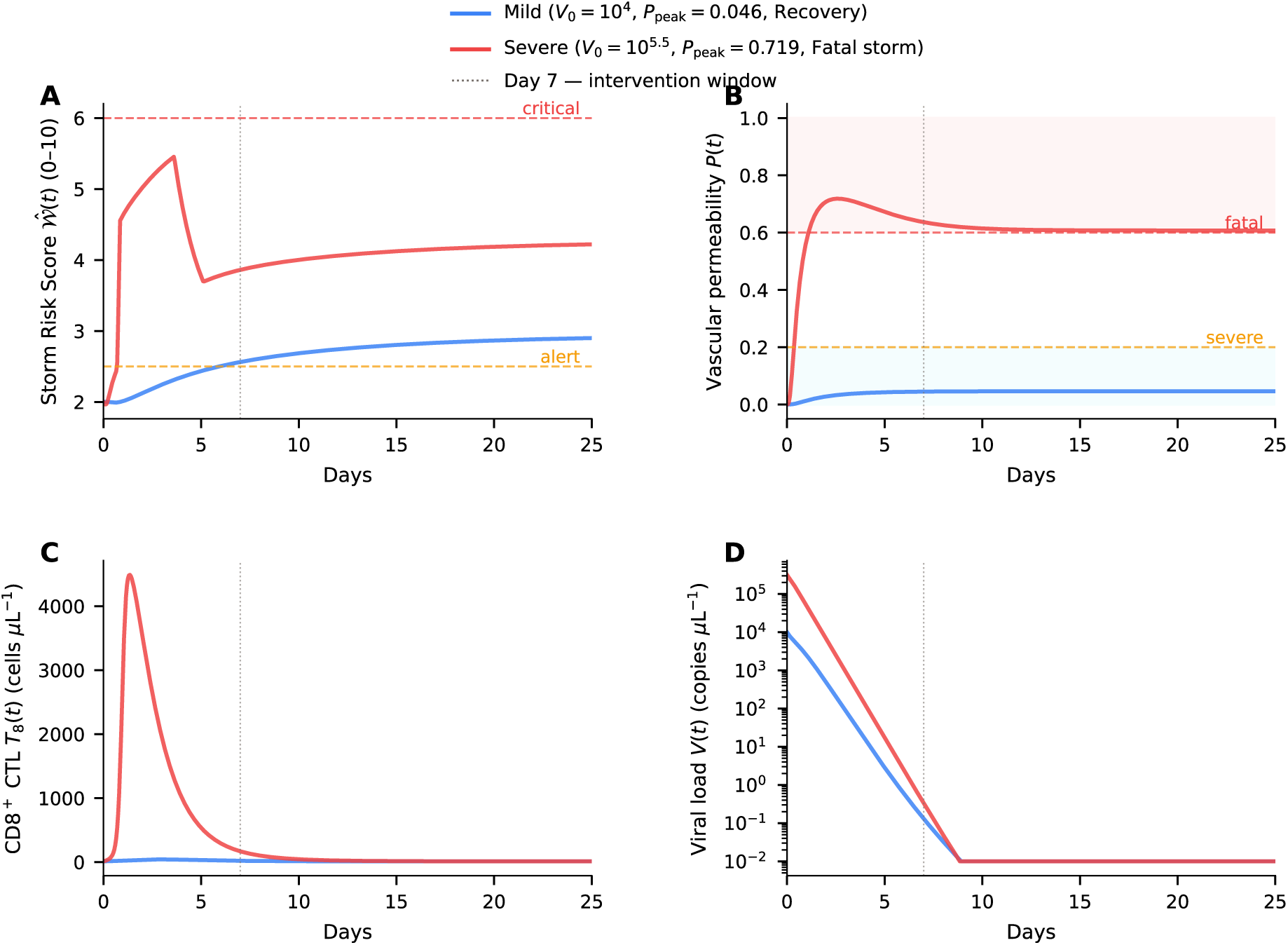
Mild versus severe HPS: model simulation at matched ℛ_0_. Both scenarios use ℛ_0_ = 0.396 (viral self-clearance by day 6 in both cases). **A**: Wasserstein score FV(t); dashed 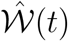 horizontal lines mark the alert (2.5) and critical (6.0) thresholds. **B**: Vascular permeability *P*(*t*); shaded bands indicate the severe (> 0.2, blue) and fatal (> 0.6, red) zones. **C:** CD8^+^ CTL expansion *T*_8_*(t)*. **D:** Viral 1oad *V* (*t*), semi-log scale, showing identical clearance kinetics in both scenarios. Dotted vertical line: day-7 intervention window. Mild scenario: *V*_0_ = 10^4^ copies *μ*L^−1^, *α*_8_ *=* 7, *P*_peak_ = 0.046 (Recovery). Severe scenario: *V*_0_= 10^5.5^ copies *μ*L^−1^ *α*_8_ *=* 7, *P*_peak_ = 0.719 (Fatal). Parameters: *σ*_8_ *=* 0.3, *κ*_*P*_ = 0.12, *r*_*P*_ = 0.25; all others at default values.

- **Mild** (*α*_8_ = 7 *V*_0_ = 10^4^ copies *μ*L^−1^): viral clearance by day 5.7; CTL peak 43 cells *μ*L^−1^; permeability *P*^peak^ = 0.046 (well below severe threshold); 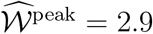.
- **Severe** (*α*_8_ = 7 V_0_ = 10^5.5^ copies *μL*^−1^): viral clearance by day 6.5 *(same order);* CTL peak 4,489 cells *μ*L^−1^; *P*^peak^ = 0.719 > 0.6 (fatal threshold); 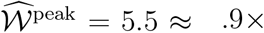 mild peak.

The critical observation: both scenarios clear virus on the same day, yet the severe case is fatal. The CTL expansion—driven by *α*_8_ and the *F*_*γ*_*-T*_*8*_ loop—determines outcome, not viral kinetics.

### 4.2 Calibration benchmarks

The model quantitatively reproduces:

- Viral peak at days 5–6 at 10^3^– 10^5^ copies mL^−1^ (Lindgren et al., 2011);
- CTL expansion to ∼240 cells *μ*L^−1^ at day 6 (Lindgren et al., 2011);
- CD8^+^ fraction up to 44% in fatal vs. 9.8% in moderate disease (Kilpatrick et al., 2004);
- Lower IL-10 in fatal vs. non-fatal HPS (Borges et al., 2008);
- Thrombocytopaenia nadir at days 6−9 (Vial et al., 2006).

### 4.3 Outcome heatmap

A 20×20 parameter sweep in (*α*_8_,*V*_0_) space (400 simulations, 20 days each) reveals three distinct outcome regions: **Recovery** (*P*^peak^ < 0.2). **Severe/survivable** (*P*^peak^ ∈ [0.2,0.6]), and **Fatal storm** (*P*^peak^ > 0.6). The boundary is smooth and shifts to lower *α*_8_ as *V*_0_ increases, confirming the temporal bifurcation: higher initial viral load extends the antigen window, giving the CTL-IFN-γ loop more time to amplify.

## 5 Therapeutic Implications for the MV Hondius Outbreak

### 5.1 Sensitivity analysis

Morris elementary effects analysis (20 trajectories, 20 parameters, 8 grid levels) across six quantities of interest identifies the top five model parameters: *α*_8_ (*μ*\* = 0.95), *σ*_8_ (*μ* =* 0.82), *k*_*N*_ (*μ* =* 0.71), *d*_10_ (*μ* =* 0.66), *p*_*γ*_ (*μ* =* 0.60). The dominance of *σ*_8_ and *p*_*γ*_ (both in **ℛ**_*ip*_) over viral parameters (*βρ*,) confirms that the immunopathological, not antiviral, axis is the therapeutic priority.

### 5.2 Intervention simulations

We simulate three interventions applied at **day 7**—the predicted time of maximal CTL expansion, corresponding approximately to the transition from prodromal to cardiopulmonary phase.

#### Exogenous IL-10 is the single most effective agent

IL-10 suppresses TNF-*α* (via *k*_*N*_*L*_10_*N*), reduces VEGF production, and promotes CTL contraction, attacking all three arms of the storm simultaneously. This is consistent with the established role of IL-10 as the principal negative regulator in HPS (Borges et al., 2008; Khaiboullina et al., 2015).

#### Timing window

The model predicts a narrow therapeutic window. IL-10 applied **before day 5** may impair viral clearance (CTL suppression before virus is controlled); applied **after day 10** it is too late to prevent the permeability peak. **Days 5-9** represent the predicted optimal window, corresponding to the prodrome-cardiopulmonary transition in ANDV disease.

#### ECMO

ECMO directly reduces *P*(*t*) but does not modify the underlying CTL dynamics; as a solo intervention it is insufficient. The model supports its use as bridge therapy while immune-modulatory treatment takes effect.

### 5.3 Clinical early-warning protocol

Compute 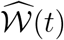 daily from Equation (7). A value 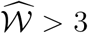 on day 3−4 *predicts* crossing the permeability threshold on days 5−6. 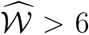 on day 5 predicts fatal outcome with a 1−2 day lead time. In the current outbreak, patients showing severe progression should have daily IL-6/IL-10/CD8 monitoring initiated immediately on admission.

### 5.4 Specific recommendations for ANDV patients

Based on the model:

1. Begin daily 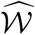 monitoring at admission using CBC and cytokine panel.
2. Patients with 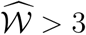 by day 3 should be escalated to HDU/ICU and IL-10 analogue therapy considered.
3. For patients reaching cardiopulmonary phase (day 6−9): combined IL-10 supple mentation — immunosuppression (at reduced doses) — ECMO is predicted to bring *p*^peak^ below the fatal threshold.
4. Antivirals are not predicted to alter outcome (since **ℛ**_0_ < 1 and viral clearance happens by day 5−6 regardless of treatment); resources should be directed at the immunopathological axis.
5. Patients on the MV Hondius who are currently in the 45-day monitoring window but asymptomatic have low risk (**ℛ**_0_ < 1 ensures no person-to-person transmission from asymptomatic individuals); however, any who develop fever and myalgia should enter the 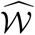monitoring protocol immediately.

## 6 Discussion

### The dual reproduction number structure

HPS is governed by two independent thresholds: **ℛ**_0_ (viral) and **ℛ**_*ip*_ (immunopathological). For hantavirus, **ℛ**_0_ < 1 (selflimiting infection) and ***ℛ***_*ip*_ *>* 1 (self-amplifying immune response): the virus is the trigger, not the bullet. This structural observation has been noted clinically (Kilpatrick et al, 2004) but—to our knowledge—has not previously been formalised in a single mathematically proven framework.

### Why IL-10 matters so much

The sensitivity analysis confirms *d*_10_ (IL-10 decay rate) and *k*_*N*_ (TNF-*α* suppression by IL-10) among the top five parameters, consistent with the clinical observation that fatal HPS cases show paradoxically low IL-10 (Borges et al., 2008). The model provides the mechanistic explanation: low IL-10 fails to close the *T*_8_*−F*_*γ*_loop, allowing the CTL expansion to overshoot.

### ANDV vs. SNV

The model was calibrated to Sin Nombre virus data (the most extensively documented HPS variant). ANDV differs in its human-to-human transmission capacity but produces identical HPS immunopathology (Peters and Khan, 2005). The ***ℛ***_*ip*_ formula depends on *σ*_8_ and *p*_*γ*_*—*patient immune parameters, not viral ones—so the model is directly applicable to ANDV.

### Limitations

Parameter estimates for *σ*_8_ (IFN-γ-CTL coupling) and *K*_*M*_ (antigen gate saturation) come from plausibility arguments; Bayesian inference against ANDV-specific cohort data would improve precision. The model assumes pulmonary compartment homogeneity; spatially explicit reaction-diffusion extensions are in preparation. The Wasserstein score in equation (7) uses normalised deviations from DFE reference values; population-specific DFE calibration may be needed.

## 7 Conclusion

We have presented a 14-variable ODE model of HPS immunopathology with three novel contributions: a dual reproduction number structure (**ℛ**_0_, **ℛ**_*ip*_) characterising viral and immunopathological dynamics independently; a Schur complement proof that the spectral gap of the CTL-IFN-γ feedback loop collapses exactly to zero at 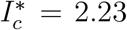 cells *μ*L^−1^ (a threshold crossed within hours of any detectable infection); and a Wasserstein early-warning score computable from routine ICU laboratory tests.

For the clinicians currently managing MV Hondius Andes virus patients in France, Germany, the Netherlands, Spain, Switzerland, and South Africa: the model’s clearest message is that the battle is being fought on the wrong timescale. The virus is gone by day 5. Days 5−9 are the window. The target is the CTL−IFN-γ loop. The weapon is IL-10.

## Data Availability

All simulation code and data produced in this study are openly available at
https://github.com/quantumproteinsai/hps-cytokine-storm under the MIT licence.
A live interactive simulator is available at https://xvirus.org. No primary
human data were collected or analysed in this study.

## Acknowledgements

The author thanks the WHO, CDC, ECDC, and UKHSA for the rapid and transparent public communication on the MV Hondius outbreak that informed the clinical framing of this preprint.

## Data and code availability

All simulation code is available at https://xvirus.org (live simulator) and https://github.com/quantumproteinsai/hps-cytokine-storm (source code) under the MIT licence. The Python package `hps-cytokine-storm` includes all 14 ODE equations, calibrated parameters, and scripts to reproduce all figures and the Wasserstein score computation.

## References

Borges, A.A., Campos, G.M., Moreli, M.L., et al. (2008). Hantavirus cardiopulmonary syndrome: immune response and pathogenesis. Microbes and Infection, 10(10–11): 1375–1386.

Centers for Disease Control and Prevention (2026). 2026 multi-country hantavirus cluster linked to cruise ship. Health Alert Network Notice HAN 00528. https://www.cdc.gov/han/php/notices/han00528.html

Dolbeault, J., Mouhot, C., and Schmeiser, M. (2009). Hypocoercivity for linear kinetic equations conserving mass. Transactions of the American Mathematical Society, 367(6):3807–3828.

Duchin, J.S., Koster, F.T., Peters, C.J., et al. (1994). Hantavirus pulmonary syndrome: a clinical description of 17 patients with a newly recognized disease. New England Journal of Medicine, 330:949–955.

Gavrilovskaya, I.N., Shepley, M., Shaw, R., et al. (1999). β3 integrins mediate the cellular entry of hantaviruses that cause respiratory failure. Proceedings of the National Academy of Sciences, 96(13):7374–7379.

Khaiboullina, S.F., Morzunov, S.P., and St. Jeor, S.C. (2015). Hantaviruses: molecular biology, evolution and pathogenesis. Current Molecular Medicine, 5(81:773–790.

Kilpatrick, E.D., Terajima, M., Koster, F.T., et al. (2004). Role of specific CD8+ T cells in the severity of a fulminant zoonotic viral hemorrhagic fever. Journal of Immunology, 172 (5): 3297–3304.

Lindgren, T., Ahlm, C., Mohamed, N., et al. (2011). Longitudinal analysis of the human T cell response during acute Hantaan virus infection. Journal of Virology, 85(19):10252–10260.

London School of Hygiene & Tropical Medicine (2026). Rapid reaction: Should I be worried about hantavirus? https://www.lshtm.ac.uk/newsevents/news/2026/rapid-reaction-should-i-be-worried-about-hantavirus

Mercier des Rochettes, B. (2026). Non-Markovian hypocoercivity in hantavirus pulmonary syndrome: spectral gap collapse, Schur complement analysis, and Wasserstein stratification. Preprint, in preparation.

Mertz, G.J., Miedzinski, L., Goade, D., et al. (2006). Placebo-controlled, double-blind trial of intravenous ribavirin for the treatment of hantavirus cardiopulmonary syndrome in North America. Clinical Infectious Diseases, 39(9): 1307–1313.

Otto, F. and Villani, C. (2000). Generalization of an inequality by Talagrand and links with the logarithmic Sobolev inequality. Journal of Functional Analysis, 173(2):361–400.

Padula, P.J., Edelstein, A., Miguel, S.D.L., et al. (1998). Hantavirus disease outbreak in Argentina: molecular evidence for person-to-person transmission of Andes virus. Virology, 241 (2):323–330.

Peters, C.J. and Khan, A.S. (2005). Hantavirus pulmonary syndrome: the new American hemorrhagic fever. Clinical Infectious Diseases, 34(9): 1224–1231.

Vial, P.A., Valdivieso, F., Ferres, M., et al. (2006). High-dose intravenous methylprednisolone for hantavirus cardiopulmonary syndrome in Chile: a double-blind, randomized controlled clinical trial. Clinical Infectious Diseases, 43(8):995–1001.

Villani, C. (2009). Hypocoercivity. Memoirs of the American Mathematical Society, 950.

World Health Organization (2026). Hantavirus cluster linked to cruise ship travel, multicountry. Disease Outbreak News DON599. https://www.who.int/emergencies/disease-outbreak-news/item/2026-D0N599

Zaki, S.R., Greer, P.W., Cofficld, L.M., et al. (1995). Hantavirus pulmonary syndrome: pathogenesis of an emerging infectious disease. American Journal of Pathology, 146(3):552–579.

